# Whole exome sequencing in the UK Biobank reveals risk gene *SLC2A1* and biological insights for major depressive disorder

**DOI:** 10.1101/2021.05.04.21256398

**Authors:** Ruoyu Tian, Tian Ge, Jimmy Z. Liu, Max Lam, Biogen Biobank team, Daniel F. Levey, Joel Gelernter, Murray B. Stein, Ellen A. Tsai, Hailiang Huang, Todd Lencz, Heiko Runz, Chia-Yen Chen

## Abstract

Nearly two hundred common-variant depression risk loci have been identified by genome-wide association studies (GWAS)^1–4^. However, the impact of rare coding variants on depression remains poorly understood. Here, we present the largest to date exome analysis of depression based on 320,356 UK Biobank participants. We show that the burden of rare disruptive coding variants in loss-of-function intolerant genes is significantly associated with depression risk. Among 30 genes with false discovery rate (FDR) <0.1, *SLC2A1*, a blood-brain barrier glucose transporter underlying GLUT1 deficiency syndrome^5–7^, reached exome-wide significance (*P*=2.96e-7). Gene-set enrichment supports neuron projection development and muscle activities^2, 3^ as implicated in depression. Integrating exomes with polygenic risk revealed additive contributions from common and rare variants to depression risk. The burden of rare disruptive coding variants for depression overlapped with that of developmental disorder, autism and schizophrenia. Our study provides novel insight into the contribution of rare coding variants on depression and genetic relationships across developmental and psychiatric disorders.

## Main

Major depressive disorder (MDD) is a common and heritable psychiatric disorder with high medical and socioeconomic burden^8, 9^. Depression GWAS have successfully identified a large number of common-variant risk loci, while the identification of rare coding variants contributing to depression risk has failed to keep pace^1–4^. Recently, large-scale exome sequencing studies of developmental and other psychiatric disorders have uncovered novel risk genes and shared genetic signals between neuropsychiatric disorders^10–15^, suggesting the promise of novel discoveries from exome analysis of depression.

To investigate the role of rare variants in depression, we analyzed exome and health data from 454,787 participants of the UK Biobank (UKB). It has been shown that the genetic architecture changes with the stringency of depression definitions in the UKB and, therefore, we identified cases and controls for each of seven previously reported depression phenotypes with different levels of strigency^16^. These included phenotypes for individuals who sought medical help for depression from either a general practitioner or a specialist (GPpsy, Psypsy); were clinically documented or self-reported as showing symptomatic depression (DepAll, EHR [electronic health record], SelfRepDep); or had one of two Composite International Diagnostic Interview (CIDI) based clinical diagnoses (lifetimeMDD, MDDRecurr) (**Table S1**). We also removed participants with self-reported substance abuse, psychotic condition or bipolar disorder from our analysis. Using UKB exome sequencing data, we annotated rare coding variants (minor allele frequency [MAF] <1e-5) by their predicted effects on the respective protein into three categories: protein-truncating variants (PTV), missense variants (further categorized by the MPC deleteriousness score^17^), and synonymous variants. We also stratified genes by pLI (probability of loss-of-function intolerance)^18, 19^ and used this to classify rare variants further. Annotated rare variants were aggregated into 11 groups across the predicted mutation severity spectrum, including 6 groups for rare variants in pLI ≥0.9 genes (PTV, MPC > 2, 2 ≥ MPC >1, 1 ≥MPC >0, other missense variants without MPC annotation and synonymous variants) and 5 groups for pLI <0.9 genes (PTV, 2 ≥MPC >1, 1 ≥MPC >0, other missense variants without MPC annotation and synonymous variants).

We first assessed the impact of exome-wide burden of rare variants on depression risk in unrelated individuals of European (EUR) descent (N=320,356). Exome-wide PTV burden significantly increased depression risk in GPpsy-, Psypsy-, SelfRepDep- and EHR-defined phenotypes (**Figure S1, Table S2**), with the most prominent associations in loss-of-function (LoF) intolerant genes (pLI ≥ 0.9) (**Figure 1a**). The strongest signal was observed in EHR-defined depression (OR=1.17, 95% CI=1.13-1.21, *P*=3.57e-18) (**Figure 1a, Table S2**). The burden of damaging missense variants (MPC>2) was significantly associated with EHR-defined depression after Bonferroni correction across all tests performed (OR=1.08, 95% CI=1.05-1.23, *P*=8.52e-6). Burden of missense variants not annotated by MPC was associated with Psypsy (**Figure 1a, Table S2**) to a lesser extent. No association was found for burden in LoF tolerant genes (pLI<0.9) (**Figure 1b**). We repeated this analysis in UKB participants of South Asian (N=7,053) and African (N=6,290) ancestries, but did not find any significant association, presumably due to limited sample sizes (**Figure S2, S3, Table S3**). Depression is more prevalent in females than males^20^. We thus conducted sex-stratified analyses for exome-wide burden. The effects of PTV and damaging missense variant burden were not statistically different between males and females (**Figure S4, Table S5, S6**). Our results demonstrate that exome-level PTV and damaging missense variant burden contribute to the risk of depression as defined by EHR in UKB.

**Figure 1.**
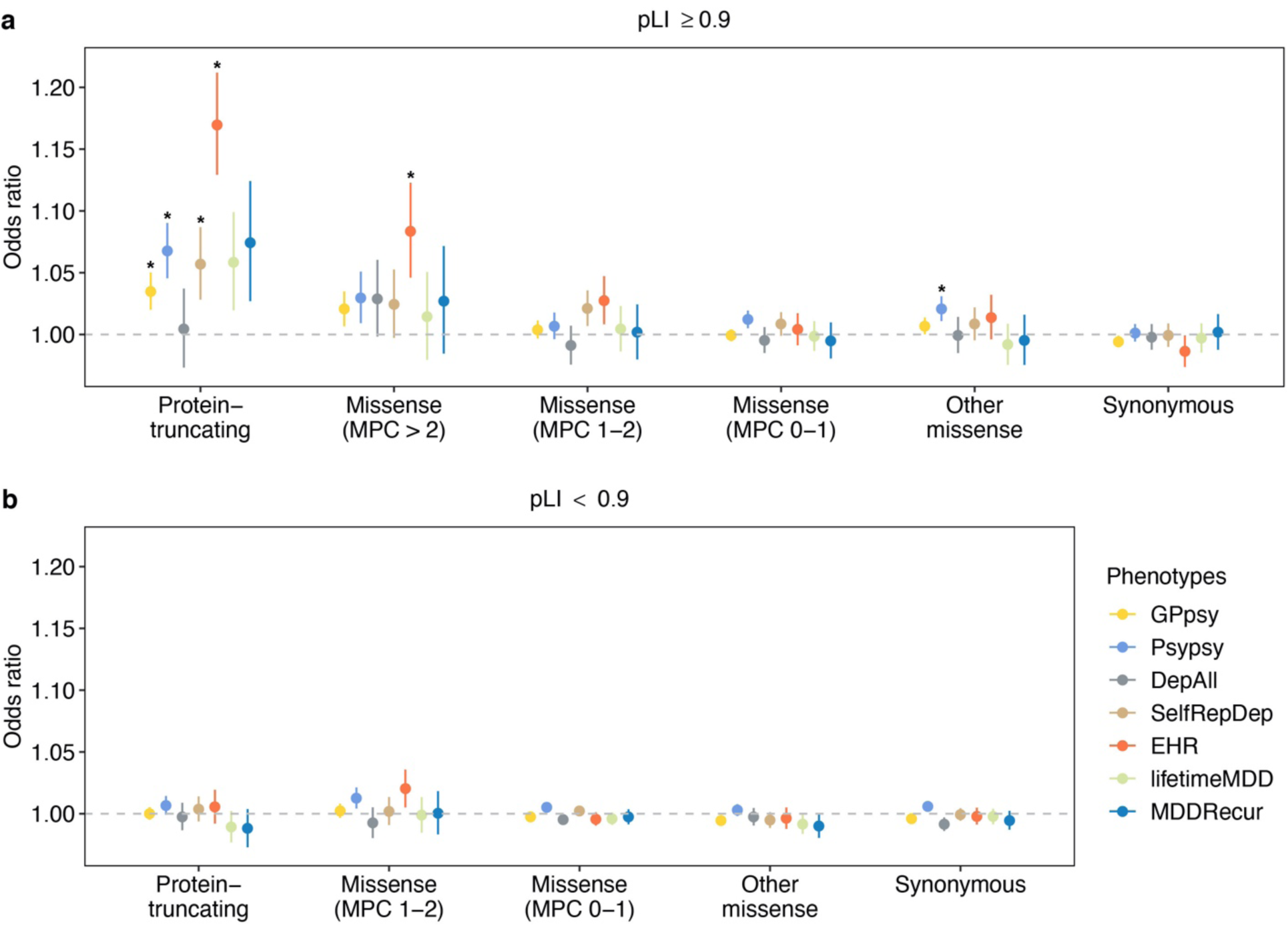
The association of rare coding variant burden with depression using seven different definitions. Y-axis is the odds ratio (OR) of the association between rare variant burden and depression risk. Protein-coding genes were stratified by pLI into (a) pLI ≥ 0.9 and (b) 0.9 > pLI genes. Rare variants were grouped by functional severity from the least to the most severe, protein-truncating, missense (MPC > 2, 2 ≥ MPC > 1, 1 ≥ MPC > 0), other missense (missense variants without MPC score annotation) and synonymous variants. Missense variants on genes (pLI < 0.9) were only annotated into two categories, 2 ≥ MPC > 1 and 1 ≥ MPC > 0. The sample size for each depression phenotype definition are as follows: GPpsy: N_cases_ = 111,712, N_controls_ = 206,617; Psypsy: N_cases_ = 36,556, N_controls_ = 282,452; DepAll: N_cases_ = 20,547, N_controls_ = 55,746); SelfRepDep: N_cases_ = 20,120, N_controls_ = 226,578; EHR: N_cases_ = 10,449, N_controls_ = 246,719; lifetimeMD: N_cases_ = 15,580, N_controls_ = 43,104; MDDRecur: N_cases_ = 9,462, N_controls_ = 43,104. The grey dashed line represents the null association (OR = 1). Each point shows the point estimate of OR from logistic regression. Bars show 95% confidence intervals (CI). *Bonferroni-adjusted significant association, *P* < 4.20e-4 (0.05/119).

Given the strongest exome PTV and damaging missense variant burden signal was found in the EHR-defined cohort (N_cases_=10,449; N_controls_=246,719), we sought to identify individual depression risk genes using the EHR-based definition of depression. We conducted 13,828 and 2,876 gene-based association tests for PTV and damaging missense burden, respectively (genes with <10 carriers were removed). The burden of damaging missense variants in *SLC2A1* (pLI=0.99, N_carriers_=52, OR=6.01, 95% CI=3.03-11.94, *P*=2.96e-7) was significantly associated with depression at an exome-wide significance level (*P*<1.74e-05), with a total of 30 genes (27 for PTV, 3 for damaging missense variant) showing false discovery rate (FDR) <0.1 (**Figure 2, Table S7, Figure S5**). SLC2A1 protein is expressed in endothelial cells of the blood-tissue barriers and facilitates transport of glucose into the brain and other tissues^21^. Mutations in *SLC2A1* impair energy supply for the brain and cause GLUT1 deficiency syndrome, characterized by infantile seizures and developmental delay^5–7^. A prior study reported increased methylation of *SLC2A1* in depression cases, although a link to gene expression has not been investigated^22^. To explore the potential impact of regulatory variants on *SLC2A1* expression, we fine-mapped *SLC2A1* blood *cis*-eQTLs^23^ by SuSiE^24^, which identified two credible sets. One of the credible sets contained two eSNPs (rs2229682 and rs11537641) and is located close to a cluster of damaging missense variants that were only observed in EHR-defined depression cases (**Figure S6**). We conducted an *SLC2A1* damaging missense variant burden phenome-wide association study (PheWAS) across 1,820 ICD10 codes and 214 quantitative traits (**Figure S7, Table S8**) to identify potential pleiotropic effects of *SLC2A1*. The only significant phenotype was major depressive disorder as defined by ICD10 code F32.9 (OR=5.40, *P*=3.38e-6), further validating the association between *SLC2A1* and depression. Taken together, these results provide strong evidence that predicted damaging missense variants in *SLC2A1* increase the risk of depression.

**Figure 2.**
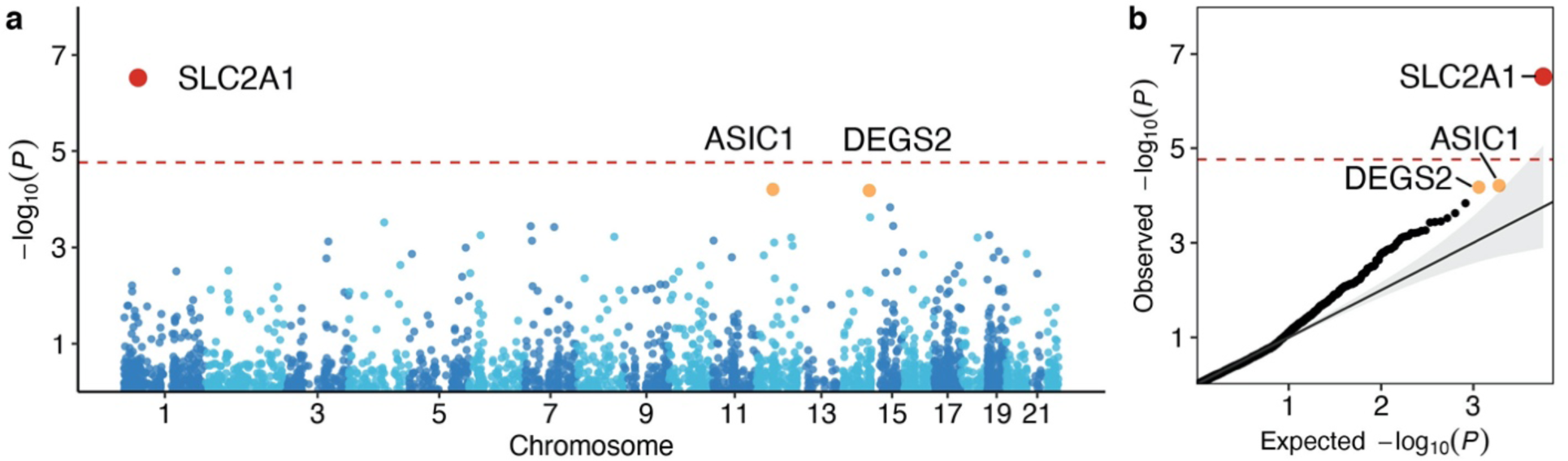
Depression gene discovery and *SLC2A1* characteristics. (a) Manhattan plots of the -log10 *P* of the association of gene-level damaging missense variant burden with EHR-defined depression. Each dot represents a gene and its genomic location is plotted on the x-axis. (b) Q-Q plots of the observed -log10 *P* of the association of gene-level damaging missense burden with EHR-defined depression against expected -log10 *P* under the null. The red dashed line is the Bonferroni significant threshold, *P* < 1.74e-5 (0.05/2,876) and the red dot represents a significant gene. The orange dots represent genes with FDR < 0.1.

Next, we aimed to functionally characterize the rare-variant burden of depression risk by self-contained gene-set analyses. Using brain expression profiles from the human protein atlas (HPA)^25^, we found significant enrichment of PTVs in brain expressed genes (OR=1.30, 95% CI=1.20–1.41, *P*=4.40e-10) compared to the baseline exome-wide signal (OR=1.03, 95% CI =1.01-1.04, *P*=7.19e-5), and the enrichment was stronger in genes with elevated brain expression relative to genes with lower brain expression specificity (**Figure 3a, Table S10**).

**Figure 3.**
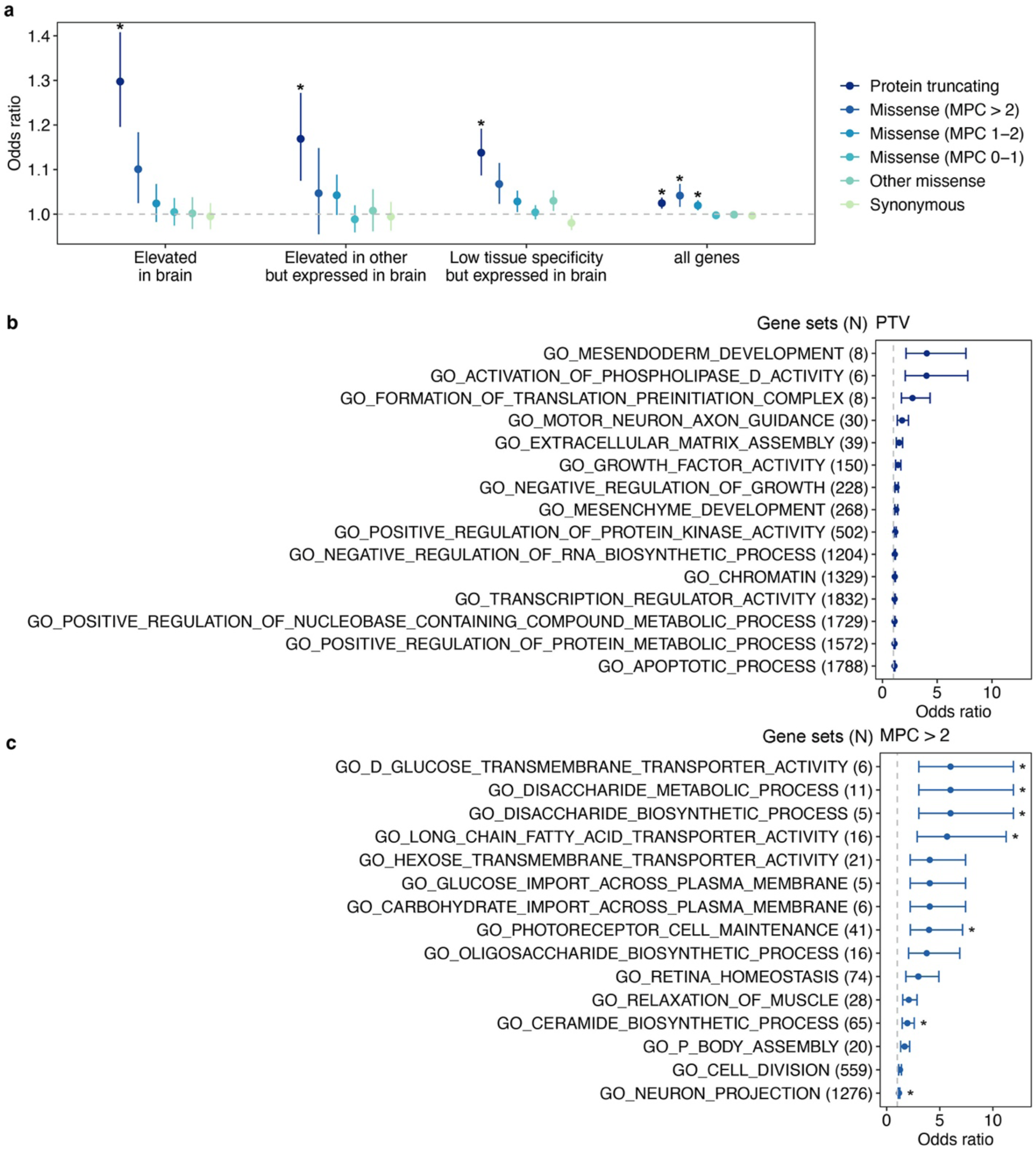
Biological insights of depression. (a) The effect of rare variants on genes stratified by brain-specific expression. We aggregated rare variants of each type (PTV, missense and synonymous) on three human brain atlas^25^ gene sets, genes with expression elevated in brain (2,587 genes), expression elevated in other tissues but expressed in brain (5,298 genes) and low tissue specificity but expressed in the brain (8,342 genes). Additionally, the exome-wide burden test served as a baseline control (“all genes”). Y-axis is the odds ratio (OR) of the association between rare variant burden on the three gene sets with depression risk. The grey dashed line represents the null (OR = 1). Each point shows odds ratio from logistic regression. Bars show 95% confidence intervals of OR. Bonferroni-adjusted significant threshold is *P* < 2.08e-3 (0.05/24). * denotes significant association. (b) Top 15 FDR-significant (FDR < 0.05) GO gene sets^26^ from PTV burden tests. (c) Top 15 FDR-significant (FDR < 0.05) GO gene sets from missense (MPC > 2) burden tests. PTV and damaging missense variants (MPC > 2) were aggregated on genes in each GO gene sets. X-axis is the odds ratio of the association between rare variant burden for each gene set (y-axis) and depression. The grey dashed line represents the null association (OR = 1). Each point shows the odds ratio from logistic regression. Bars show 95% confidence. *Bonferroni-significant association at *P* < 4.87e-6 (0.05/10,266) for (b) and *P* < 5.27e-6 (0.05/9,491) for (c), respectively.

Depression-relevant biological pathways, cell types and tissues have been implicated in common variant analyses^3, 4^. To investigate the potential biological mechanism underlying rare-variant associations, we performed gene-set based burden analyses on 10,271 gene sets, including biological processes (N=7,573), cellular components (N=1,001) and molecular function (N=1,697)^26^. In total, we identified 18 and 60 unique gene sets (FDR<0.05) for PTV and damaging missense variant burden, respectively (**Figure 3b, c, Table S11a, b**). These gene sets converged on neuron projection development, muscle activities and chromatin remodeling, as implicated in GWAS pathway analyses, stratified LD score regression (LDSC) and animal studies^2, 3, 27^. Fifteen of the 60 gene sets identified through missense burden were driven by the genetic risk in *SLC2A1*, while only 4 gene sets survived multiple testing correction (FDR<0.05) after excluding *SLC2A1* from the damaging missense burden (**Table S11c**)^28^. Finally, we queried whether rare variant associations inform the development of depression medications and drug repurposing. We identified 207 genetic targets of 63 FDA approved antidepressants (e.g. activator, agonist, antagonist, binder, blocker, inhibitor, ligand and modulator) (**Table S12a, b, d**) from the DGldb browser (4.2.0)^29^. There were no significant associations with variants with PTV, 2≥MPC >1, 1≥MPC> 0 and synonymous variants in the genetic targets of approved FDA therapies, but there was a significant association of the burden of missense variants not annotated by MPC with depression risk (OR=1.12, 95% CI=1.04-1.22, *P*=5.07e-3) (**Figure S8, Table S12c**). To explore drug repurposing opportunities, 35 approved or investigational drugs, which served as binder, blocker, and antibody and inhibitor of 8 FDR<0.1 genes (*SLC2A1*, *ASIC1*, *TSHZ2*, *LRRC59*, *PCSK9*, *CD38*, *CLCN3* and *RTN4*) were identified in DGldb (**Table S12d**). These findings might inform and support the discovery and development of new depression drugs and drug repurposing.

While depression GWAS and our analyses presented here show that both common and rare coding variants contribute to depression risk, their relative contributions remain unclear. To examine the polygenic background of depression risk in individuals from UKB, we performed a meta-analysis (N_cases_=157,304, N_controls_=576,282) of the depression GWAS from the Psychiatric Genomics Consortium (PGC)^2^, Million Veteran Program (MVP)^4^, and FinnGen (Release 6) to create a discovery GWAS (**Table S13**), which has no sample overlap with UKB. We then calculated a polygenic risk score (PRS) for each UKB participant using PRS-CS^30^ and the 1000 Genomes EUR samples as the reference panel, and classified each individual by their carrier status of PTV and damaging missense variants. As shown in **Figure 4a**, in both carriers and non-carriers of damaging rare variants, the prevalence of depression increased with higher PRS, while given the same polygenic risk, carriers of damaging rare variants had increased risk of depression relative to non-carriers. To quantify the relative contributions of common and rare genetic components to depression, we fitted a joint logistic regression to PRS and the carrier status of PTV and damaging missense variants. In the EHR cohort, common variants (PRS) (OR per SD change in PRS =1.34, 95% CI=1.31-1.37, *P*=4.77e-184), PTV (OR per risk allele =1.16, 95% CI=1.12-1.21, *P*=4.26e-17) and damaging missense variants (OR per risk allele =1.07, 95% CI=1.04-1.11, *P*=6.26e-5) explained 2.51%, 0.22% and 0.06% of the total phenotypic variation on the liability scale^31^, respectively (**Table S14**). Notably, PRS explained 9-fold greater variance than rare variants. There was no significant interaction between damaging coding variant carrier status and PRS for all depression definitions (*P*>0.25) (**Table S14, Figure S9**), suggesting additive contributions from PRS and rare variants to depression risk.

**Figure 4.**
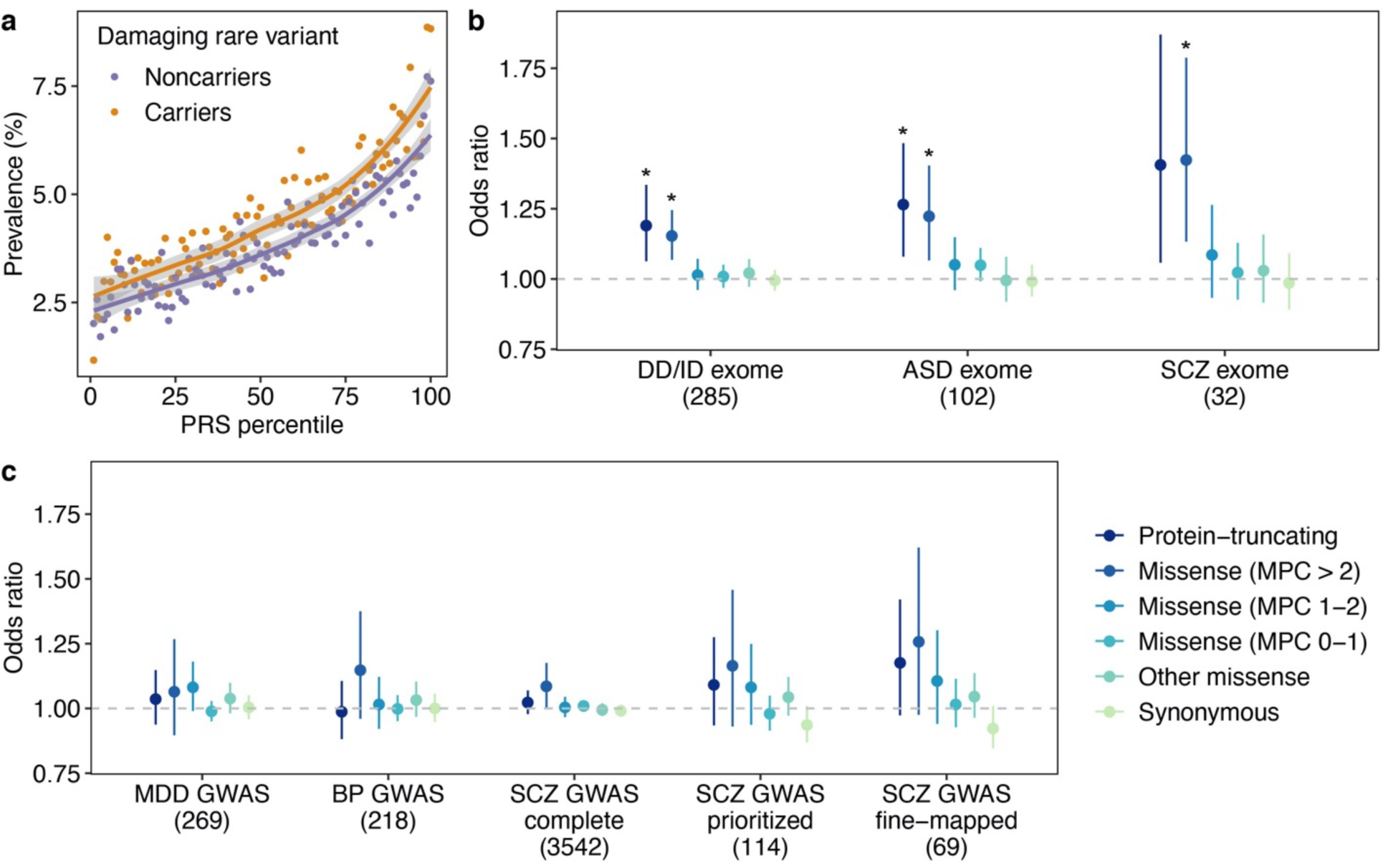
Additive contributions from rare and common variants to depression risk. (a) Scatter plot of the prevalence of EHR-defined depression against PRS percentile for PTV or damaging missense variant on LoF intolerant genes for carriers and noN_carriers_. The lines are fitted by loess regression and grey shading corresponds to the 95% confidence interval of the fit. (b) The effect of rare variants on psychiatric and neurodevelopmental disease associated genes identified from exome analysis and (c) genes identified from GWAS. We aggregated rare variants of each type (PTV, missense and synonymous) on 8 disease gene sets. For exome identified risk genes, we obtained 102 (FDR < 0.1), 285 (Bonferroni significant) and 32 (FDR < 0.05) putative risk genes discovered from exome analyses of autism (ASD)^10^, developmental disorder (DD/ID)^14^ and schizophrenia (SCZ)^12^, respectively. For depression (MDD) GWAS, we obtained 269 genes identified by meta-analysis^3^. For bipolar disorder (BP) GWAS genes, we obtained 218 protein-coding risk genes positionally mapped from 30 GWAS loci^32^. For SCZ, we acquired the 3,542 complete positionally mapped genes (“SCZ GWAS complete”), 114 prioritized protein-coding genes (“SCZ GWAS prioritized”) and 69 fine-mapped genes from the largest meta-analysis by PGC phase 3 (“SCZ GWAS fine-mapped”)^33^. Y-axis is the odds ratio of the association between rare variant burden on each gene set with depression risk. The grey dashed line represents the null association (OR = 1). Each point shows the odds ratio from logistic regression. Bars show 95% confidence intervals. No association was Bonferroni-adjusted significant (*P* < 1.04e-3=0.05/48). *FDR < 0.05.

Lastly, we investigated the impact of rare coding variant burden in genes identified by prior GWAS or exome sequencing studies of depression related diseases. Rare coding variant burden in MDD, schizophrenia or bipolar disorder GWAS genes was not associated with depression (**Figure 4c, Table S16**). In addition, these was no depression GWAS signal in the *SLC2A1* locus^4^ (**Figure S10**), suggesting independent contributions from common and rare variants to depression risk. In contrast, genetic risk derived from exome studies were shared between depression and developmental disorders, autism spectrum disorder and schizophrenia (**Figure 4b, Table S16**), supporting the convergence of genetic risk from rare coding variants in psychiatric and developmental disorders.

In summary, we present the largest depression exome sequencing study to date. Collectively, disruptive coding variant burden in LoF intolerant genes increased depression risk. Gene burden analysis identified 30 genes at FDR<0.1, among which *SLC2A1* reached exome-wide significance. Our analyses recapitulated biological pathways in neuron projection development and muscle activities identified by prior common-variant GWAS^2, 3^. Joint analysis of genetic variants across the allele frequency spectrum revealed additive contributions from common and rare variants to depression risk. Furthermore, rare variant genetic risk was shared between depression and other psychiatric and developmental disorders. These results provide novel insights and expand our understanding of the genetic basis of depression.

## Methods

### The UK Biobank and whole-exome sequencing

The UK Biobank is a large prospective population-based study with over half a million participants recruited across the UK^34^. Phenotypic data collected from each participant includes survey measures, electronic health records, self-reported health information and other biological measurements^35^. The participants have diverse genetic ancestries and overrepresented familial relatedness^35^.

Whole-exome sequencing (WES) data from UK Biobank participants was generated by the Regeneron Genetics Center (RGC) as part of a collaboration between AbbVie, Alnylam Pharmaceuticals, AstraZeneca, Biogen, Bristol-Myers Squibb, Pfizer, Regeneron and Takeda. The WES production and quality control (QC) is described in detail in Van Hout et al^36^. As of November 2020, we obtained QC passed WES data (“*Goldilock*s” set) from 454,787 samples in the UK Biobank.

### Variant annotation

We annotated variants identified through WES by Variant Effect Predictor (VEP) v96^37^ with genome build GRCh38. Variants annotated as stop-gained, splice site disruptive and frameshift variants were further assessed Loss-Of-Function Transcript Effect Estimator (LOFTEE)^19^, a VEP plugin. LOFTEE implements a set of filters to remove variants that are unlikely to be disruptive. Those variants labeled as “low-confidence” were filtered out, and we kept variants labeled as “high-confidence”. Variants annotated as missense variants were then annotate by MPC score^17^, which prioritized damaging missense variants. All predicted variants were all mapped to GENCODE^38^ canonical transcripts. In total, we identified 649,321 predicted rare PTVs, 5,431,793 missense variants and 3,060,387 synonymous variants with minor allele frequency <10^-5^.

### Phenotyping of depression

Out of all 502,524 UK Biobank samples, we first removed 2,256 individuals with self-reported substance abuse (code 1408, 1409 and 1410 in data field 20002), self-reported manic or psychotic condition (code 1291 in data field 20002), bipolar I disorder and bipolar II disorder (code 1,2 in derived data field 20126). We then followed the seven definitions of depression described in Cai et al^16^, including two broad definitions (“GPpsy” and “Psypsy”), a symptom-based definition (“DepAll”), a self-reported definition (“SelfRepDep”), a medical-record-based definition (“EHR”), and two CIDI-based definitions (lifetimeMDD” and “MDDRecur”). The former five definitions only required a minimal number of questions to identify depression cases (minimal phenotyping), while the latter two were closer to clinical diagnosis of depression based on Composite International Diagnostic Interview Short Form (CIDI-SF) but were only available for the individuals who participated in the UK Biobank online mental health follow-up.

### Sample filtering and population assignment

We restricted our analyses to 407,139 unrelated individuals and removed 1,804 individuals whose reported gender differed from genetic sex or who had sex chromosome aneuploidies.

We also removed 133 individuals withdrawn (as of August 24, 2020) from the UK Biobank. To identify UK Biobank samples from different populations for analysis, we performed population assignment based on population structure derived using principal component analysis (PCA) with 1000 Genomes Project (1KG) reference samples (N_sample_ = 2,504) from 5 major population groups: East Asian [EAS], European [EUR], African [AFR], American [AMR], and South Asian [SAS]. We first performed quality control on the 1KG genotype data by retaining only the SNPs on autosomes with minor allele frequency (MAF) > 1% and removed SNPs located in known long-range LD regions (chr6: 25-35Mb; chr8: 7-13Mb). We also removed 1 sample from each pair of related samples (greater than second degree) in 1KG. We merged the UK biobank imputed genotype data that was filtered to include imputation quality INFO > 0.8 and MAF > 1% with the 1KG genotype data. We performed LD-pruning at R^2^ = 0.2 with a 500 kb window. We then computed principal components (PCs) using the LD-pruned SNPs in 1KG sample and derived projected PCs of UK Biobank samples using the SNP-wise PC loadings from 1KG samples. Using the 5 major population labels of 1KG samples as the reference, we trained a random forest model with top 6 PCs to classify UK Biobank samples into 1KG population groups. We assigned UK Biobank samples into one of the 5 populations defined with 1KG reference based on a predicted probability for a specific population group > 0.8. We identified 1,609 EAS samples, 458,197 EUR samples, 8,406 AFR samples, 9,224 SAS samples, 1,085 AMR samples and 8,874 samples without explicit population assignment. Due to the small sample sizes, we did not further analyze samples of EAS and AMR ancestry. We also excluded subjects without an explicit population assignment. After initial population assignment, we performed three rounds of within population PCA for AFR, EUR and SAS samples to identify remaining population outliers, each time removing samples with any of the top 10 PCs that was more than 5 SD away from the sample average. We used the in-sample PCs derived after outlier removal in subsequent analyses. We kept individuals with depression case-control status and passed sequencing QC within AFR, EUR and SAS population groups for analysis.

### Whole-exome and gene-level burden test

We grouped protein coding genes by pLI (v2.1.1)^18, 19^ into LoF intolerant (pLI ≥ 0.9) set and LoF tolerant (pLI < 0.9) set. We annotated rare variants by functional consequences into three types, protein-truncating, missense and synonymous. Missense variants were further annotated by MPC score^17^ and stratified into 4 groups by predicted outcome severity, MPC > 2, 2 ≥ MPC > 1, 1 ≥ MPC > 0 and others (referring to those missense variants without MPC annotation). In total, we have 11 sets of variants: PTV, MPC > 2, MPC > 2, 2 ≥ MPC > 1, 1 ≥ MPC > 0, other missense and synonymous variants in pLI ≥ 0.9 genes; PTV, 2 ≥ MPC > 1, 1 ≥ MPC > 0, other missense and synonymous variants in pLI < 0.9 genes. Note that missense variants on pLI < 0.9 genes were not annotated into MPC > 2 category. Rare alleles of the same variant category on each gene were aggregated into gene-level burden. The summation of the burden on genes in each gene set was the whole-exome burden.

For the whole-exome burden test, we applied logistic regression (“glm” function in R) by fitting whole-exome burden to depression case-control status as the binary response. In the model, we controlled for population structure with top 20 PCs, mean centered age, sex, mean centered age^2^, mean centered age × sex, mean centered age^2^ × sex. Additionally, we included the 22 assessment centers as categorical covariates (**Table S4**). We performed 119 logistic regressions across 7 curated phenotype definitions and 17 variant sets. We defined a significant threshold *P* < 4.20e-4 (0.05/119) for the whole-exome burden tests.

For gene-level burden test, we fitted a Firth’s logistic regression by regressing case-control status on the burden on each gene, a binary variable denotating rare allele carrying status. We restricted to PTV and damaging missense (MPC>2) burden and EHR-defined depression for gene-level burden test, based on significant association in the whole-exome burden tests. We included the same covariates as described above. We excluded genes with less than 10 carriers for PTV or damaging missense burden. In total, 16,704 association tests were conducted, including 13,828 tests for PTV and 2,876 tests for damaging missense variant. Exome-wide significance was *P* < 2.99e-6 (0.05/16,704) or FDR < 0.05.

### Sex-specific analyses

Depression has roughly double prevalence in females compared to males and we sought to examine the potential sex-specific effect of rare variant burden. We first tested the exome-wide burden association with depression in males and females in a logistic regression controlling for mean centered age, mean centered age^2^ and top 20 PCs for all 11 variant categories in the EHR-derived cohort (N of tests = 33). We also tested the association for protein-truncating and damaging missense variant burden in LoF intolerant genes for the other 6 phenotype definitions (N of tests = 36). Significance threshold was *P* < 7.25e-4 (0.05/69) or FDR < 0.05 for the sex-specific analysis. We further tested if the number of rare variants per sample is different in affected males and affected females, or in control males and in control females. Two-sided Poisson exact test was performed across 7 phenotype definitions and 2 comparisons (affected female against affected male; control female against male) for PTV and damaging missense variants. In total, there were 28 independent tests and the Bonferroni significant level was *P* < 1.79e-3 (0.05/28).

### *SLC2A1 cis*-eQTL fine-mapping

Blood *SLC2A1 cis*-eQTL full summary statistics were acquired from eQTLGen^23^ browser. For the *cis*-eQTL fine-mapping approach, we implemented a summary statistics based Sum of Single Effects (SuSiE)^24^. LD reference was precalculated from 1000 Genome European samples. The maximum number of causal variants was set as 10. After fitting SuSiE regression, we examined the posterior inclusion probability for each variant in the 95% credible sets.

### Phenome-wide association study (PheWAS)

We performed a damaging missense variant burden of *SLC2A1* PheWAS across 2,034 binary and quantitative phenotypes. Each binary phenotype was derived from an ICD10 code in the UK Biobank and was mapped to a Phecode. We excluded phenotypes with less than 100 cases for binary phenotypes, which yielded 1,820 binary phenotypes in the PheWAS. We took a two-step approach to first test all gene-phenotype pairs by logistic regression and then performed Firth’s logistic regression for those gene-phenotype pairs passed significant threshold (*P* < 0.05). For quantitative phenotypes, we excluded phenotypes with fewer than 100 observations and phenotypes with less than 12 distinct values. For each phenotype, we removed individuals with phenotype value > 5 SDs from the sample mean. Burden testing was performed using linear regression on both the raw and inverse rank-based normal transformed quantitative phenotypes. We controlled for top 20 PCs, mean centered age, sex, mean centered age^2^, mean centered age × sex, mean centered age^2^ × sex and assessment centers in the PheWAS. We defined phenome-wide significant thresholds as *P* < 2.46e-5 (0.05/2,034) and FDR < 0.05.

### Gene set enrichment analyses

We tested if damaging rare variant burden were enriched in specific functional gene sets, gene ontology (GO)^26^, the human brain proteome^25^, antidepressant interacted genes^29^, major depressive disorder GWAS risk genes^3^ and other neuropsychiatric and neurodevelopmental disease associated genes^10–14, 32, 33^. We applied logistic regressions by fitting an individual disease status on the number of rare variants in a given gene set the individual carried, controlling for 20 PCs, mean centered age, sex, mean centered age^2^, mean centered age × sex and mean centered age^2^ × sex.

#### Gene ontology

We acquired 10,271 GO gene sets from MSigDB v7.2^26^, including biological process (N = 7,573), cellular component (N = 1,001) and molecular function (N = 1,697), which are derived from the Biological Process Ontology by the Gene Ontology Consortium^39, 40^. We applied Firth’s logistic regression^41^ for testing the hypothesis. For variant category, we only tested PTV and damaging missense variant with EHR-defined depression given the results from our whole-exome burden analysis. We defined Bonferroni-adjusted significant threshold for PTV and damaging missense variant as *P* < 4.87e-6 (0.05/10,266) and *P* < 5.27e-6 (0.05/9,491), respectively, or FDR < 0.05.

#### Brain specific expression

The Human Protein Atlas (HPA) – Brain Atlas^25^ integrated 1,710 RNA-seq samples cross 23 human brain regions from GTEx, cap analysis of gene expression (CAGE) and HPA. In the Brain Atlas, 16,227 genes were kept for analysis after normalization and filtering. Those genes were then categorized by their relative expression in brain and other tissues: expression elevated in brain (2,587 genes), expression elevated in other tissues but expressed in brain (5,298 genes) and expression was not tissue specific but expressed in brain (8,342 genes) **(Table S9)**. We applied logistic regressions to test for the association of all 6 variant categories cross the three gene sets. We defined a significance threshold *P* < 2.78e-3 (0.05/18).

#### Antidepressants interacted genes

We obtained drug-gene interactions (updated in April 13, 2021) from the DGldb browser (4.2.0)^29^, a database collection of drug-gene interactions and druggable genes from publications and web sources. There are four categories of FDA approved antidepressants, tricyclic antidepressants (TCAs), selective serotonin antidepressants (SSRIs), serotonin and norepinephrine reuptake inhibitors (SNRIs) and other (moclobemide). And there are 21, 9 and 33 drugs belonging to TCAs, SSRIs and SNRIs, respectively **(Table S12a).** Antidepressant interacted genes were extracted for each drug from browser, and unique genes were kept. Finally, there were 207 genes in total, served as the drug-gene interacted gene set. We performed logistic regressions on 6 types of variants on this gene set. We defined a significance threshold *P* < 8.33e-3 (0.05/6) or FDR < 0.05. To identify drug that can be repurposed for depression treatment, we also extracted drugs that interacted with 30 FDR < 0.1 depression risk genes (**Table S7**).

#### Neuropsychiatric and neurodevelopmental diseases associated risk genes

To examine the genetic risk of rare variants in genes identified through common variants in GWAS for depression and other psychiatric disorders, we tested 269 genes depression genes^3^, 218 bipolar disorder genes^32^ and 3,542 positionally mapped genes, 114 prioritized protein-coding genes and 69 fine-mapped genes for schizophrenia^33^, all identified by GWAS meta-analysis (**Table S15**). We also tested if depression shares rare genetic risk variants with other neuropsychiatric diseases and neurodevelopmental disorder. We obtained 102 (FDR < 0.1), 285 (Bonferroni significant) and 32 (FDR < 0.05) putative risk genes discovered from up-to-date whole-exome analyses of autism^11^, neurodevelopmental disorder^14^ and schizophrenia^12^, respectively. In total, there were 8 groups of disease associated genes. We conducted logistic regressions to test for disease risk association of the six types of variants in each gene set. We defined a significance threshold *P* < 1.04e-3 (0.05/48) or FDR < 0.05.

### Polygenic risk score (PRS) analysis

#### Meta-analysis

We meta-analyzed three GWAS of depression: the meta-analysis by Psychiatric Genomics Consortium (PGC)^2^ without participants from the UK Biobank or 23andme; GWAS on individuals with European ancestry from Million Veteran Program (MVP) cohort^4^; and depression GWAS from FinnGen Release 6.

Quality control (QC) pipeline of each above summary statistics underwent the following steps if information was available: 1. Remove duplicate and ambiguous SNPs, and SNPs without rsID; 2. Remove SNPs with minor allele frequency (MAF) < 0.01. We used PLINK 1.90 beta^42^ to perform an inverse-weighted fixed-effects meta-analysis of the three summary statistics. SNPs appeared in two or more studies were included in the meta-analysis. SNP heritability and LD score regression intercept were computed by LDSC v1.0.1^43^. SNP heritability on the observed scale was transformed to heritability on the liability scale^31^, where population prevalence K was set to 0.15. LD score regression intercept was used for evaluating genomic inflation for each study.

#### UK Biobank genetic data

The genome-wide genotyping was performed for all UK Biobank participants and imputed using the Haplotype Reference Consortium (HRC)^44^ and UK10K + 1000 Genomes^45^ reference panels, resulting in a total of more than 90 million variants. We carried out QC steps on the genotyping data by filtering out variants with imputation quality score less than 0.8, or variants with MAF less than 0.001 by PLINK 2.00 alpha^42^. We performed the PRS analysis in EUR samples only due to restricted sample size in AFR and SAS samples.

#### PRS calculation

We applied polygenic risk scores-continuous shrinkage (PRS-CS)^30^ to estimate the effect sizes of genetic markers. LD reference panel was precomputed using 1000 Genomes Project phase 3 samples with European ancestry (available at https://github.com/getian107/PRScs). Global shrinkage parameter phi was set to be 0.01 since depression is a highly polygenic trait. PRS of each chromosome for each individual in the validation set was computed by the “--score” function in PLINK 2.00 alpha^42^, a linear combination of genotypes weighted by effect size estimates. The final PRS was then summed across chromosome 1 to 22.

#### PRS predictive performance evaluation

To access the predictive performance of PRS, we computed and compared Cox & Snell pseudo *R*^2^ for each phenotype with the following the null model (1) and the full model (2):

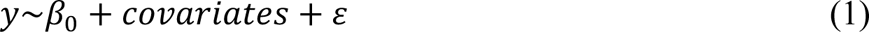

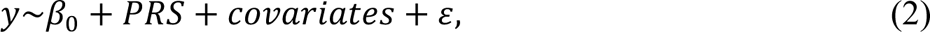

where *y* is the phenotypic binary response, *β*_0_ is the intercept, *covariates* are 20 PCs, mean centered age, sex, mean centered age^2^, mean centered age × sex and mean centered age^2^ × sex and *ε* is the random error. The partial *R*^2^ on the observed scale for PRS was estimated with the full and null generalized linear model with the same set up as above. *R*^2^ on the observed scale was then transformed to liability scale^31^. Moreover, to compare variance explained by each variance component, PRS, PTV and damaging missense variant, we also computed Cox & Snell pseudo *R*^2^, *R*^2^ on the observed scale and the liability scale for PTV and damaging missense variant by replacing the variable *PRS* with the tested term in full models. Finally, we tested for the interaction effect between PRS and rare variant in a logistic regression:

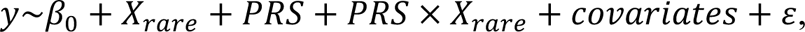

where *X*_*rare*_ is a binary variable denoting an individual carrying a protein-truncating variant or a damaging missense variant.

## Data availability

All phenotypic and genotypic data for the UK Biobank are available to researchers under data access request from the UK Biobank data access process (https://www.ukbiobank.ac.uk/enable-your-research/register). 454,787 whole exome sequencing data is not publicly available yet.

Depression GWAS summary statistics from FinnGen Release 6 is not publicly available. Meta-analysis of depression by PGC (without UK Biobank and 23andme participants) is available at https://www.med.unc.edu/pgc/download-results/mdd/. Summary statistics of GWAS on individuals with European ancestry from Million Veteran Program (MVP) cohort was obtained through MVP Project Proposal MVP200097.

pLoF Metrics is available at https://storage.googleapis.com/gcp-public-data--gnomad/release/2.1.1/constraint/gnomad.v2.1.1.lof_metrics.by_gene.txt.bgz. eQTLGen *cis*-eQTL summary statistics is available at https://www.eqtlgen.org/. Human protein atlas is available at https://www.proteinatlas.org/humanproteome/brain/human+brain. Drug gene interaction database is available at https://www.dgidb.org/.

## Code availability

Software used for analysis was PRS-CS (https://github.com/getian107/PRScs), PLINK1.90b (https://www.cog-genomics.org/plink/), PLINK2.00a (https://www.cog-genomics.org/plink/2.0) and LD Score regression (https://github.com/bulik/ldsc).

## Data Availability

All phenotypic and genotypic data for the UK Biobank are available to researchers under data access request from the UK Biobank data access process (https://www.ukbiobank.ac.uk/enable-your-research/register). 454,787 whole exome sequencing data is not publicly available yet.
Depression GWAS summary statistics from FinnGen Release 6 is not publicly available. Meta-analysis of depression by PGC (without UK Biobank and 23andme participants) is available at https://www.med.unc.edu/pgc/download-results/mdd/. Summary statistics of GWAS on individuals with European ancestry from Million Veteran Program (MVP) cohort was obtained through MVP Project Proposal MVP200097.
pLoF Metrics is available at https://storage.googleapis.com/gcp-public-data--gnomad/release/2.1.1/constraint/gnomad.v2.1.1.lof_metrics.by_gene.txt.bgz. eQTLGen cis-eQTL summary statistics is available at https://www.eqtlgen.org/. Human protein atlas is available at https://www.proteinatlas.org/humanproteome/brain/human+brain. Drug gene interaction database is available at https://www.dgidb.org/.

https://www.ukbiobank.ac.uk/enable-your-research/register

https://www.med.unc.edu/pgc/download-results/mdd/

https://storage.googleapis.com/gcp-public-data--gnomad/release/2.1.1/constraint/gnomad.v2.1.1.lof_metrics.by_gene.txt.bgz

https://www.eqtlgen.org/

https://www.proteinatlas.org/humanproteome/brain/human+brain

https://www.dgidb.org/

## Acknowledgements

We thank all the participants and researchers of the UK Biobank and FinnGen. We thank Million Veteran Program (MVP) for kindly providing the GWAS summary results of meta-analysis of depression. We thank Sally John for critically revising the manuscript. We thank the Biogen Biobank team for initiation of the UK Biobank whole exome sequencing project and their technical supports and scientific contributions.

## Author contributions

C.C. and H.R. conceived and supervised the study. R.T. and C.C. performed the analyses. R.T. wrote the manuscript. T.G., J.Z.L., M.L., D.L., J.G., M.B.S., E.A.T., H.H., T.L., H.R. and C.C. critically revised the paper. All authors reviewed and approved the final version of the manuscript.

## Supplementary figures

**Figure S1.** The exome-wide association of rare variants with seven definitions of depression in European ancestry samples.

**Figure S2.** The association of rare variants with seven definitions of depression in African ancestry samples.

**Figure S3.** The association of rare variants with seven definitions of depression in South Asian ancestry samples.

**Figure S4.** Sex-stratified association of rare coding variant burden with depression.

**Figure S5.** Manhattan plot and Q-Q plot of risk gene discovery from PTV burden test.

**Figure S6.** The regional plot of *SLC2A1 cis*-eQTLs from eQTLGen^23^.

**Figure S7.** PheWAS of the burden of damaging missense variants in *SLC2A1*.

**Figure S8.** The effect of rare variants on antidepressants interacted genes.

**Figure S9.** Prevalence of depression for PRS percentile in PTV or damaging missense variant carriers and noN_carriers_.

**Figure S10.** Regional plot of major depressive disorder (MDD) GWAS^4^ *SLC2A1* locus.

## Supplementary tables

**Table S1.** Case-Control sample sizes of depression definitions cross populations. Samples sizes in subjects with (a) European, (b) African and (c) South Asian ancestries.

**Table S2.** Exome-wide burden test statistics in EUR. Summary statistics of each regression analysis for each phenotype-rare-variant pair. “pheno”: phenotype; “BETA”: coefficient of burden; “SE”: standard error; “OR”: odds ratio; “OR_lower”: the lower bound of 95% confidence interval of odds ratio estimate; “OR_upper”: the upper bound of 95% confidence interval of odds ratio estimate; “P”: *P*; “type”: type of rare variants (“ptv”: protein-truncating variant; “MPC2”: MPC > 2; “MPC1”: MPC: 1 – 2; “MPC0”: MPC > 1; “other_MS”: missense variants without MPC annotation; “syn”: synonymous variant); “pLI”: gene is LoF tolerant (“tol”), intolerant (“intol”) or all genes (“0-1”).

**Table S3.** Exome-wide burden test statistics in (a) AFR and (b) SAS. Summary statistics of each regression analysis for each phenotype-rare-variant pair. “pheno”: phenotype; “BETA”: coefficient of burden; “SE”: standard error; “OR”: odds ratio; “OR_lower”: the lower bound of 95% confidence interval of odds ratio estimate; “OR_upper”: the upper bound of 95% confidence interval of odds ratio estimate; “P”: *P*; “type”: type of rare variants (“ptv”: protein-truncating variant; “MPC2”: MPC > 2; “MPC1”: MPC: 1 – 2; “MPC0”: MPC > 1; “other_MS”: missense variants without MPC annotation; “syn”: synonymous variant); “pLI”: gene is LoF tolerant (“tol”) or intolerant (“intol”).

**Table S4.** Exome-exome burden test statistics in EUR (with assessment center as a covariate). Summary statistics of each regression analysis for each phenotype-rare-variant pair. “pheno”: phenotype; “BETA”: coefficient of burden; “SE”: standard error; “OR”: odds ratio; “OR_lower”: the lower bound of 95% confidence interval of odds ratio estimate; “OR_upper”: the upper bound of 95% confidence interval of odds ratio estimate; “P”: *P*; “type”: type of rare variants (“ptv”: protein-truncating variant; “MPC2”: MPC > 2; “MPC1”: MPC: 1 – 2; “MPC0”: MPC > 1; “other_MS”: missense variants without MPC annotation; “syn”: synonymous variant); “pLI”: gene is LoF tolerant (“tol”) or intolerant (“intol”).

**Table S5.** Sex-stratified exome-wide burden test statistics. (a) Test statistics for EHR across all types of rare variants; (b) Test statistics for GPpsy, Psypsy, DepAll, SelfRepDep, lifetimeMDD and MDDRecur on PTV and damaging missense variant burden of LoF intolerant genes. “pheno”: phenotype; “BETA”: coefficient of burden; “SE”: standard error; “OR”: odds ratio; “OR_lower”: the lower bound of 95% confidence interval of odds ratio estimate; “OR_upper”: the upper bound of 95% confidence interval of odds ratio estimate; “P”: *P*; “type”: type of rare variants (“ptv”: protein-truncating variant; “MPC2”: MPC > 2; “MPC1”: MPC: 1 – 2; “MPC0”: MPC > 1; “other_MS”: missense variants without MPC annotation; “syn”: synonymous variant); “pLI”: gene is LoF tolerant (“tol”) or intolerant (“intol”); “sex”: “all” denotes all sex; “0” denotes female; and “1” denotes male.

**Table S6.** Sex-stratified two-sided Poisson exact test in (a) cases and in (b) controls. Column names in (a), “case_F”: number of female cases; “case_F_ptv”: number of PTV in female cases; “case_F_ms”: number of damaging missense variants (MPC > 2) in female cases; “case_M”: number of male cases; “case_M_ptv”: number of PTV in male cases; “case_M_ms”: number of damaging missense variants (MPC > 2) in male cases; “P_ptv_case”: *P* of the Poisson-exact test for the number of PTV per sample in female cases against the one in male cases; “P_ms_case”: *P* of the Poisson-exact test for the number of damaging missense variants (MPC > 2) per sample in female cases against the one in male cases. Column names in (b), replace “case” with “ctrl” (control).

**Table S7.** Depression genes identified in gene-based analyses for PTV and damaging missense variant burden (FDR < 0.1). Genes identified from (**a**) damaging missense variant burden and (**b**) PTV burden with EHR-defined depression. OR: odds ratio; 95% CI: 95% confidence interval. FDR were adjusted for 13,828 tests for damaging missense variant burden and 2,876 tests for PTV burden.

**Table S8.** PheWAS of damaging missense variant burden on *SLC2A1* summary statistics. Column names: “long_pheno”: complete phenotype name; “pheno”: phenotype name; “p”: *P*; “SE”: standard error; “test_type”: regression type used in the association test, including linear regression, logistic regression and Firth’s logistic regression; “category”: the phecode category that the phenotype belongs to.

**Table S9.** The human brain atlas genes. (a) 2,587 genes, expression elevated in brain; (b) 5,298 genes, expression elevated in other tissues but expressed in brain; (c) 8,342 genes, expression was not tissue specific but expressed in brain.

**Table S10.** Burden test statistics in the human brain atlas gene sets. The summary statistics of associations between rare variants burden on gene sets, elevated expression in brain (“brain”), elevated expression in other tissues but expressed in brain (“other”), expressed in brain but not tissue specific (“nospe”) and whole genes (“all_genes”). “pheno”: phenotype; “BETA”: coefficient of burden; “SE”: standard error; “OR”: odds ratio; “OR_lower”: the lower bound of 95% confidence interval of odds ratio estimate; “OR_upper”: the upper bound of 95% confidence interval of odds ratio estimate; “P”: *P*; “type”: type of rare variants (“ptv”: protein-truncating variant; “MPC2”: MPC > 2; “MPC1”: MPC: 1 – 2; “MPC0”: MPC > 1; “other_MS”: missense variants without MPC annotation; “syn”: synonymous variant).

**Table S11.** Significant (FDR < 0.05) GO gene sets and burden test statistics for (a) PTV and (b) damaging missense variant. (c) Significant GO gene sets and burden test statistics for damaging missense variant after excluding damaging missense variant burden in *SLC2A1*. “N_GENE”: the number of genes in each gene set. “OR”: odds ratio; “OR_lower”: the lower bound of 95% confidence interval of odds ratio estimate; “OR_upper”: the upper bound of 95% confidence interval of odds ratio estimate; “P”: *P.* Bonferroni-adjusted significant threshold for (a) and (b) are *P* < 4.87e-6 (0.05/10,266) and *P* < 5.27e-6 (0.05/9,491), respectively.

**Table S12.** Antidepressants interacted genes, depression risk genes interacted drugs and burden test statistics. (a) FDA approved antidepressants and the category each drug belongs to, including tricyclic antidepressants (TCAs), selective serotonin antidepressants (SSRIs), serotonin and norepinephrine reuptake inhibitors (SNRIs) and other (moclobemide). (b) 207 genes interacted with antidepressants listed in (a). (c) The summary statistics of associations between rare variants burden on the interacted genes with depression risk. “pheno”: phenotype; “BETA”: coefficient of burden; “SE”: standard error; “OR”: odds ratio; “OR_lower”: the lower bound of 95% confidence interval of odds ratio estimate; “OR_upper”: the upper bound of 95% confidence interval of odds ratio estimate; “P”: *P*; “type”: type of rare variants (“ptv”: protein-truncating variant; “MPC2”: MPC > 2; “MPC1”: MPC: 1 – 2; “MPC0”: MPC > 1; “other_MS”: missense variants without MPC annotation; “syn”: synonymous variant). Significant threshold *P* < 8.33e-3 (0.05/6) or FDR < 0.05. (d) Antidepressant-gene associations acquired from the DGldb browser. (e) Depression risk genes (FDR < 0.1) interacted drugs and associations acquired from the DGldb browser.

**Table S13.** Sample sizes, number of significant genomic risk loci, heritability, LD score intercept and mean chi-squared of depression GWAS used in the meta-analysis.

**Table S14.** Pseudo *R*^2^, *R*^2^ on the liability scale of PRS, PTV and damaging missense variant (MPC > 2), and *P* value for *PRS* × *X*_*rare*_ of each phenotypic definition.

**Table S15.** Neuropsychiatric and neurodevelopmental disease associated genes. “BP”: bipolar disorder; “ASD”: autism spectrum disorder; “DDID”: neurodevelopmental disorder; “SCZ”: schizophrenia; “MDD”: major depressive disorder. For schizophrenia GWAS genes: “GWAS_all”: 3,542 complete positionally mapped genes; “GWAS_priority”: 114 prioritized protein-coding genes; “GWAS”: 69 fine-mapped genes.

**Table S16.** Burden test statistics in neuropsychiatric and neurodevelopmental disease associated genes. “BP”: bipolar disorder; “ASD”: autism spectrum disorder; “DDID”: neurodevelopmental disorder. For schizophrenia GWAS gene set, “scz_gwas_all”: 3,542 complete positionally mapped genes; “scz_gwas_pri”: 114 prioritized protein-coding genes; “scz_gwas”: 69 fine-mapped genes. “pheno”: phenotype; “BETA”: coefficient of burden; “SE”: standard error; “OR”: odds ratio; “OR_lower”: the lower bound of 95% confidence interval of odds ratio estimate; “OR_upper”: the upper bound of 95% confidence interval of odds ratio estimate; “P”: *P*; “type”: type of rare variants (“ptv”: protein-truncating variant; “MPC2”: MPC > 2; “MPC1”: MPC: 1 – 2; “MPC0”: MPC > 1; “other_MS”: missense variants without MPC annotation; “syn”: synonymous variant). Significant threshold *P* < 1.04e-3 (0.05/48) or FDR < 0.05.

