## Supplementary Information for "Whole exome sequencing in the UK Biobank reveals risk gene *SLC2A1* and biological insights for major depressive disorder"

**Supplementary materials**

**Consortium information**

**Biogen Biobank team**

Steering team: Ellen Tsai, Christopher D. Whelan, Paola Bronson, David Sexton, Sally John, Heiko Runz;

Data management team: Eric Marshall, Mehool Patel, Saranya Duraisamy, Timothy Swan;

Extended Scientific team: Dennis Baird, Chia-Yen Chen, Susan Eaton, Jake Gagnon, Feng Gao, Cynthia Gubbels, Yunfeng Huang, Varant Kupelian, Kejie Li, Dawei Liu, Stephanie Loomis, Helen McLaughlin, Adele Mitchell, Nilanjana Sadhu, Benjamin Sun, Ruoyu Tian

### Supplementary figures

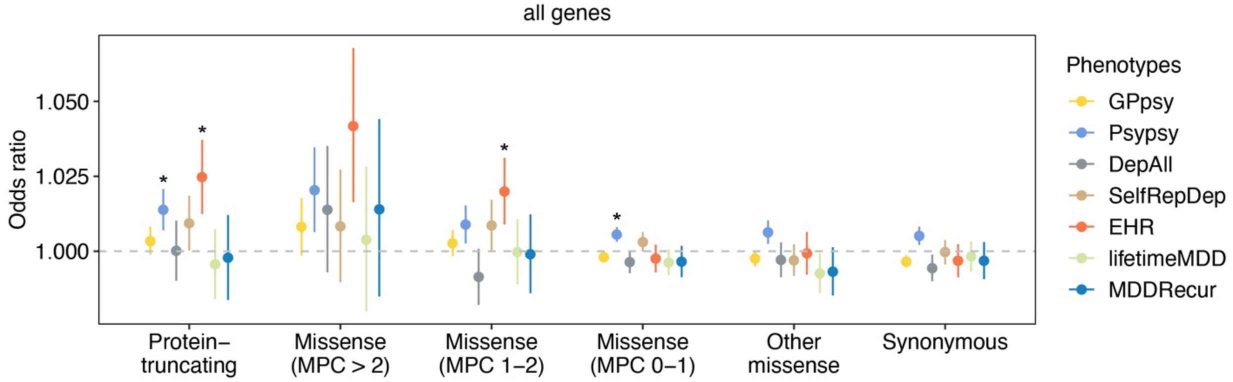


**Figure S1. The exome-wide association of rare variants with seven definitions of depression in European ancestry samples.** Y-axis is the odds ratio (OR) of the association between rare variant burden and depression risk. Rare variants were grouped by functional severity from the least to the most severe, protein-truncating, missense (MPC > 2, 2 ≥ MPC > 1, 1 ≥ MPC > 0), other missense (missense variants without MPC score annotation) and synonymous variants. The grey dashed line represents the null association (OR = 1). Each point shows the point estimate of OR from logistic regression. Bars show 95% confidence intervals (CI). * denotes Bonferroni-adjusted significant *P* < 4.20e-4 (0.05/119).

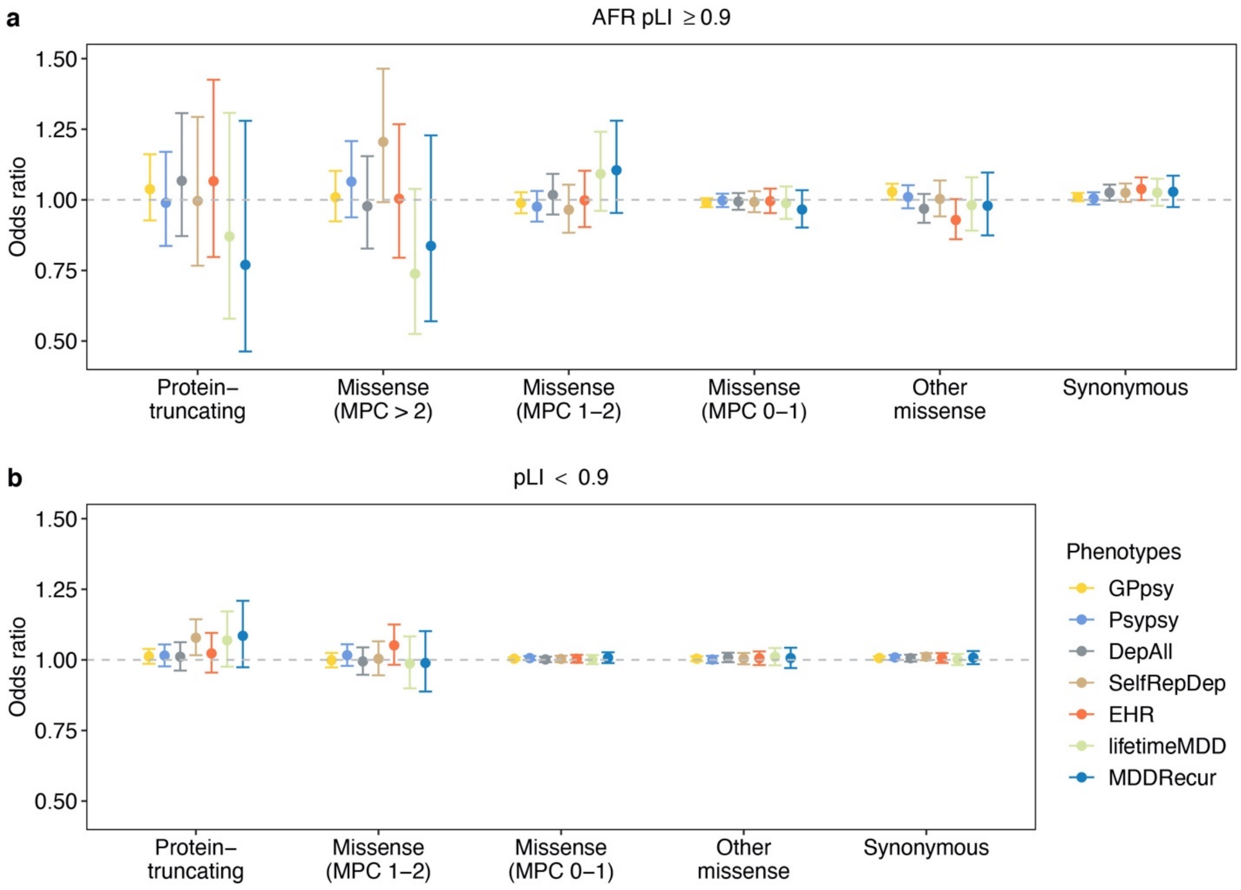


**Figure S2. The association of rare variants with seven definitions of depression in African ancestry samples.** Y-axis is the odds ratio (OR) of the association between rare variant burden and depression risk. Protein-coding genes were stratified by pLI, into (**a**) pLI ≥ 0.9 and (**b**) 0.9 > pLI genes. Rare variants were grouped by functional severity from the least to the most severe, protein-truncating, missense (MPC > 2, 2 ≥ MPC > 1, 1 ≥ MPC > 0), other missense (missense variants without MPC score annotation) and synonymous variants. Missense variants on genes (pLI < 0.9) were only annotated into 2 ≥ MPC > 1 or 1 ≥ MPC > 0. The grey dashed line represents the null association (OR = 1). Each point shows the point estimate of OR from logistic regression. Bars show 95% confidence intervals (CI). Bonferroni-adjusted significant threshold is $P < 6.49\text{e-}4$ (0.05/77).

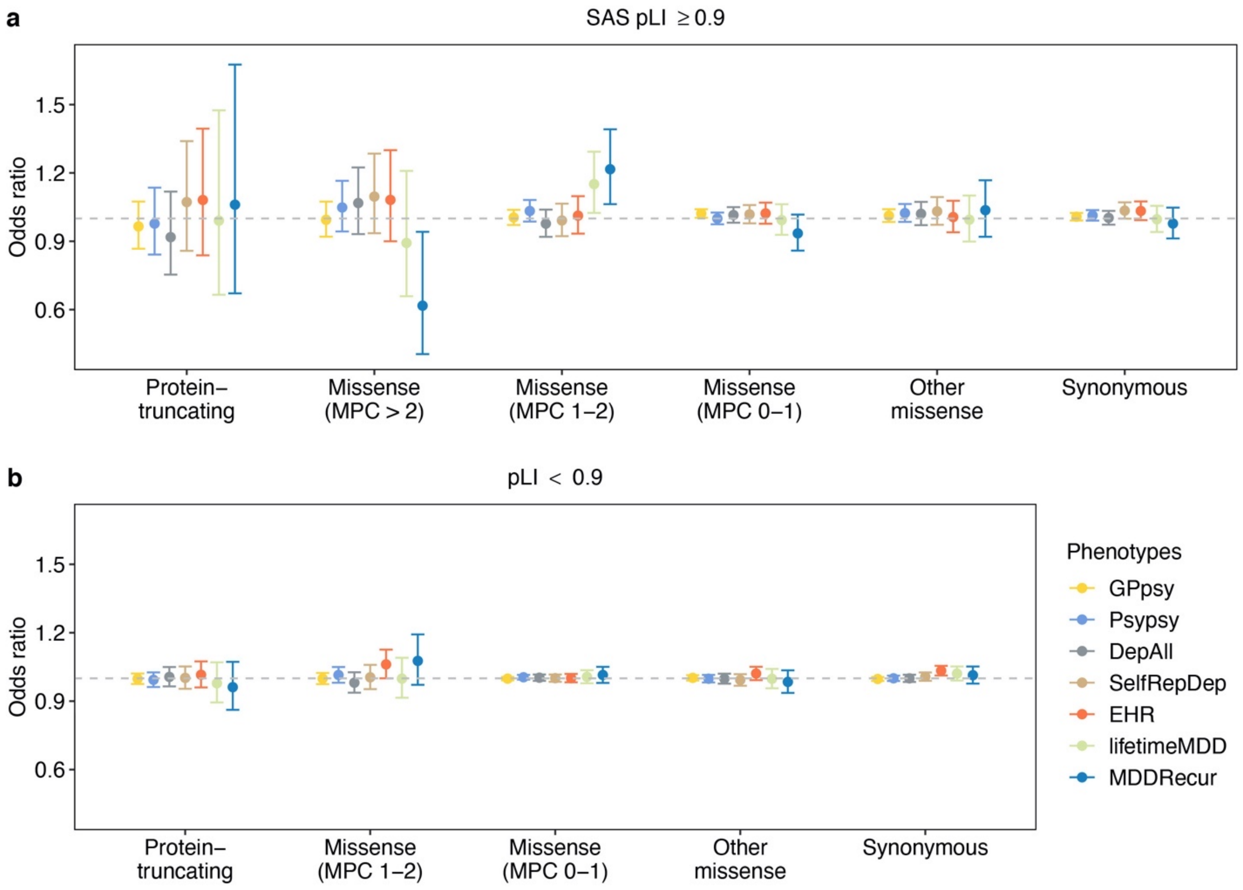


**Figure S3. The association of rare variants with seven definitions of depression in South Asian ancestry samples.** Y-axis is the odds ratio (OR) of the association between rare variant burden and depression risk. Protein-coding genes were stratified by pLI, into (**a**) pLI ≥ 0.9 and (**b**) 0.9 > pLI genes. Rare variants were grouped by functional severity from the least to the most severe, protein-truncating, missense (MPC > 2, 2 ≥ MPC > 1, 1 ≥ MPC > 0), other missense (missense variants without MPC score annotation) and synonymous variants. Missense variants on genes (pLI < 0.9) were only annotated into 2 ≥ MPC > 1 or 1 ≥ MPC > 0. The grey dashed line represents the null association (OR = 1). Each point shows the point estimate of OR from logistic regression. Bars show 95% confidence intervals (CI). Bonferroni-adjusted significant threshold is $P < 6.49e\text{-}4$ (0.05/77).

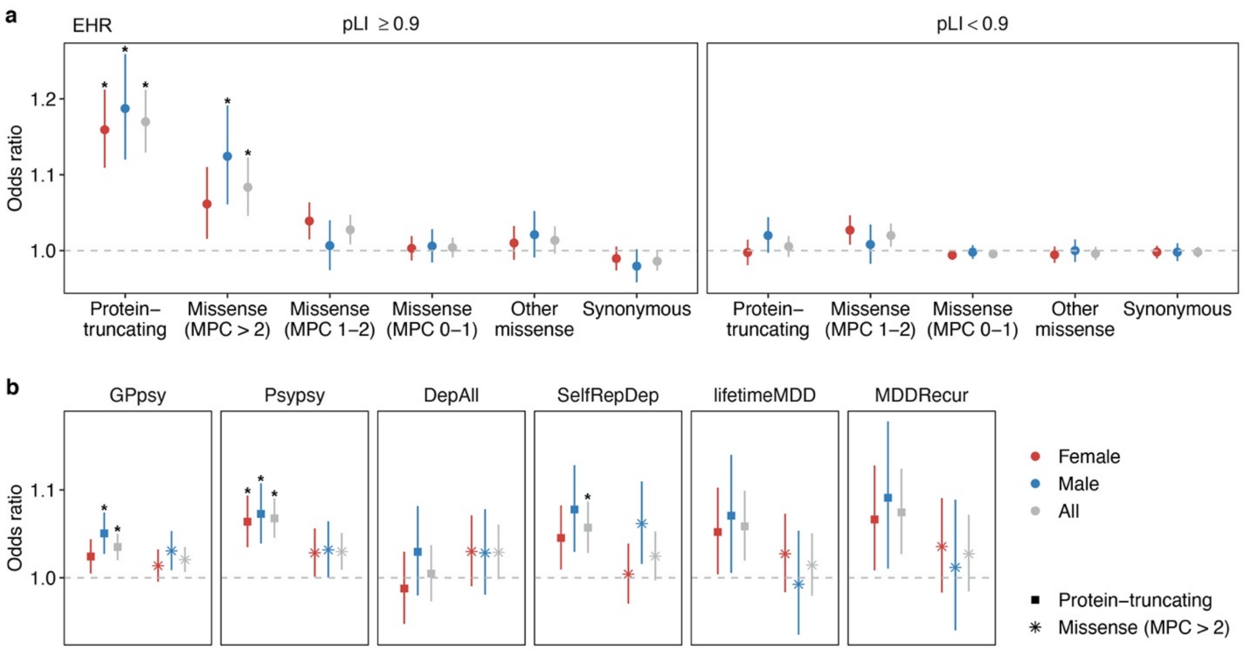


**Figure S4. Sex-stratified association of rare coding variant burden with depression.** Y-axis on each panel is the odds ratio (OR) of the association between rare variant burden and depression risk. (**a**) The effect of 11 groups of rare variants on EHR-defined depression risk in female, male and all subjects. (**b**) The effect of PTV and damaging missense variants on genes (pLI ≥ 0.9) on GPpsy, Psypsy, DepAll, SelfRepDep, lifetimeMDD and MDDRecur in female, male and all subjects. The grey dashed line represents the null association (OR = 1). Each point shows the OR from logistic regression. Bars show 95% confidence intervals (CI) of the point estimate. *Bonferroni-adjusted significant association for $P < 7.25e\text{-}4$ (0.05/69).

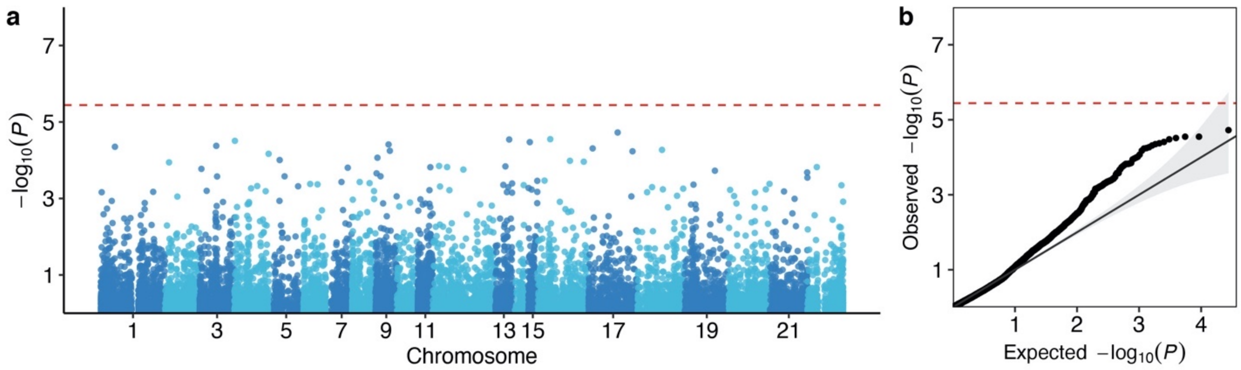


**Figure S5. Manhattan plot and Q-Q plot of risk gene discovery from PTV burden test.** (**a**) Manhattan plots of the -$\log_{10}$ *P* of the association of gene-based PTV burden with EHR-defined depression. (**b**) Q-Q plots of the observed the -$\log_{10}$ *P* of the association of gene-based PTV burden with EHR-defined depression against expected -$\log_{10}$ *P* from the null. Each dot represents a gene and its genomic location is plotted on the x-axis. The red dashed line is the Bonferroni significant threshold ($P$ < 3.62e-6 = 0.05/13,828).

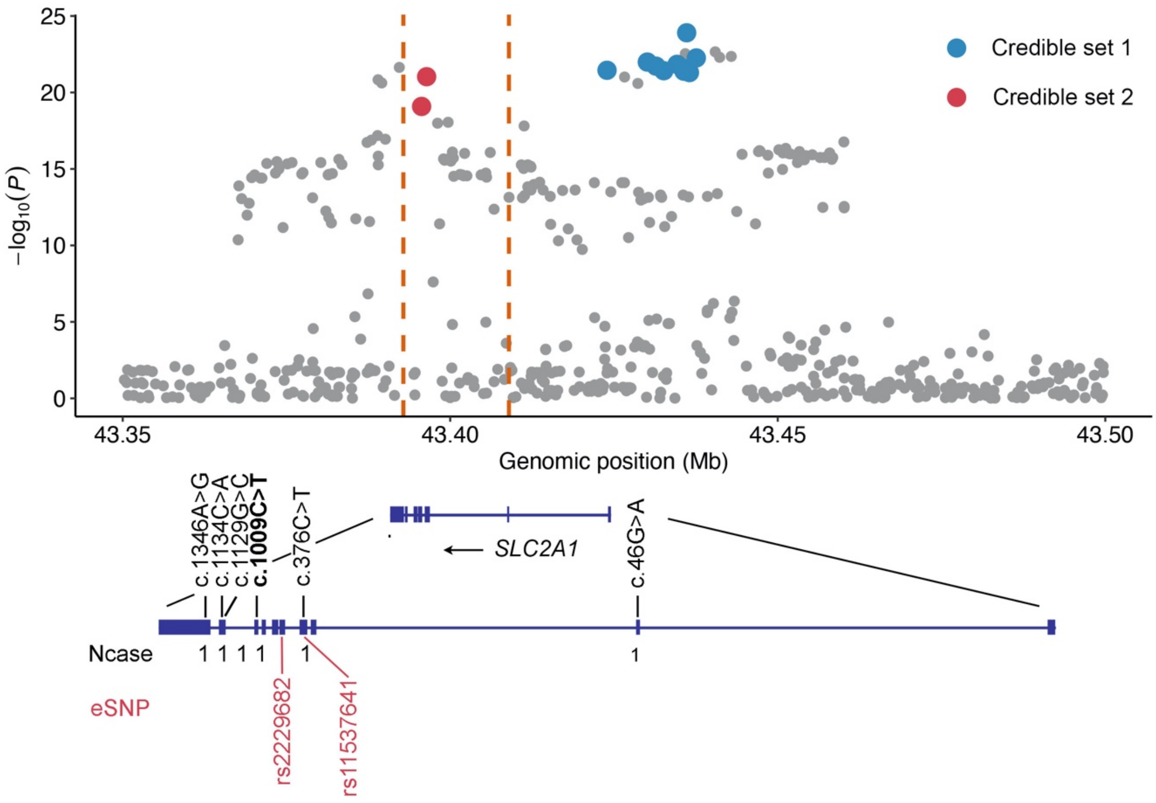


**Figure S6. The regional plot of *SLC2A1 cis*-eQTLs from eQTLGen[1]**. Each variant is plotted by its genomic location (x-axis) against -$\log_{10}$ *P* from *cis*-eQTL association (y-axis). The blue and red dots correspond to fine-mapped variants in credible set 1 and credible set 2, respectively. The orange dashed lines mark the positional range of the damaging missense variants found in depression cases. *SLC2A1* gene model is shown at the bottom with damaging missense variants in cases are labeled on top of the gene model. Variant c.1009C>T (rs80359818) in bold is pathogenic for GLUT1 deficiency syndrome reported in ClinVar (OMIM #138140). Two causal eSNPs are shown in red.

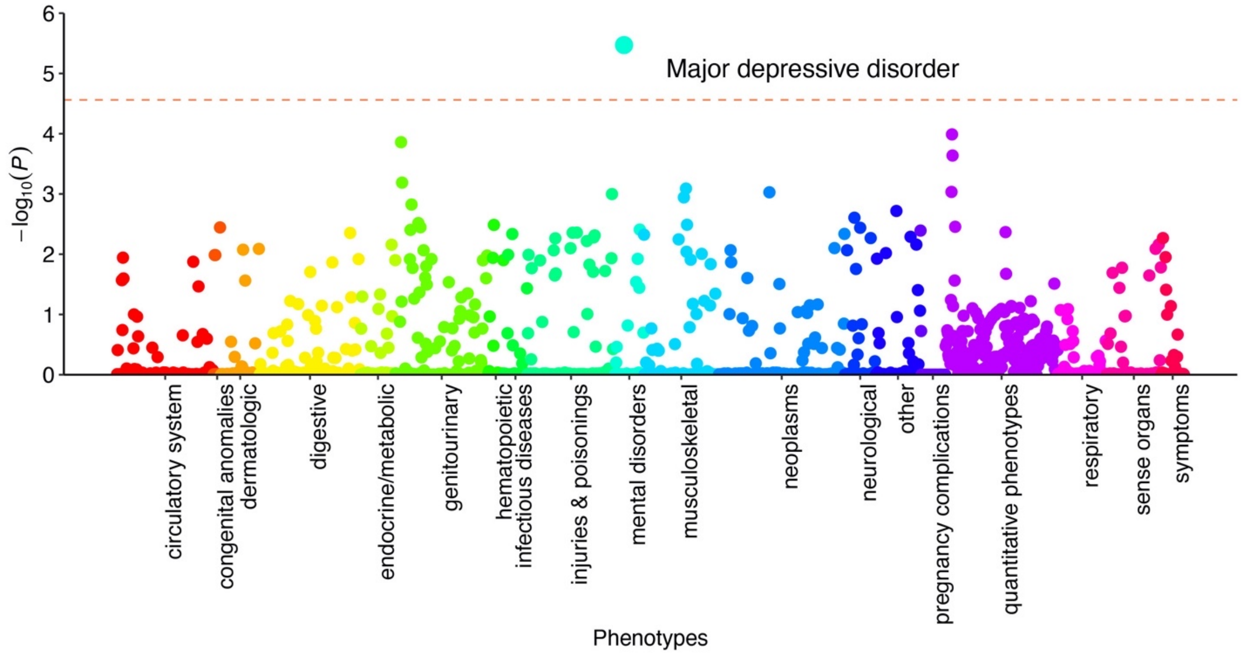


**Figure S7. PheWAS of the burden of damaging missense variants in *SLC2A1*.** Manhattan plot of the -$\log_{10}$ *P* of the association of the burden of damaging missense variants in *SLC2A1* with ICD10-defined phenotypes and quantitative traits. Each ICD10-defined phenotype mapped to a Phecode cross 18 categories on the x-axis. 214 quantitative phenotypes were groups into one label. The orange dashed line is the Bonferroni significant threshold (*P* < 2.46e-5).

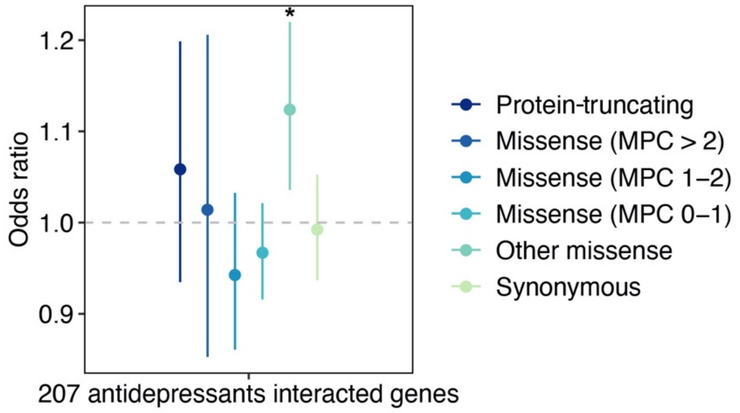


**Figure S8. The effect of rare variants on antidepressants interacted genes.** We aggregated rare variants of each type (PTV, missense and synonymous) on 207 antidepressants interacted genes. Y-axis is the odds ratio of the association between rare variant burden with depression risk. The grey dashed line represents the null association. Each point shows the point estimate of odds ratio of the logistic regression. Bars show 95% confidence intervals of the point estimate. * shows Bonferroni-adjusted significant association ($P$ < 8.3e-3 = 0.05/6).

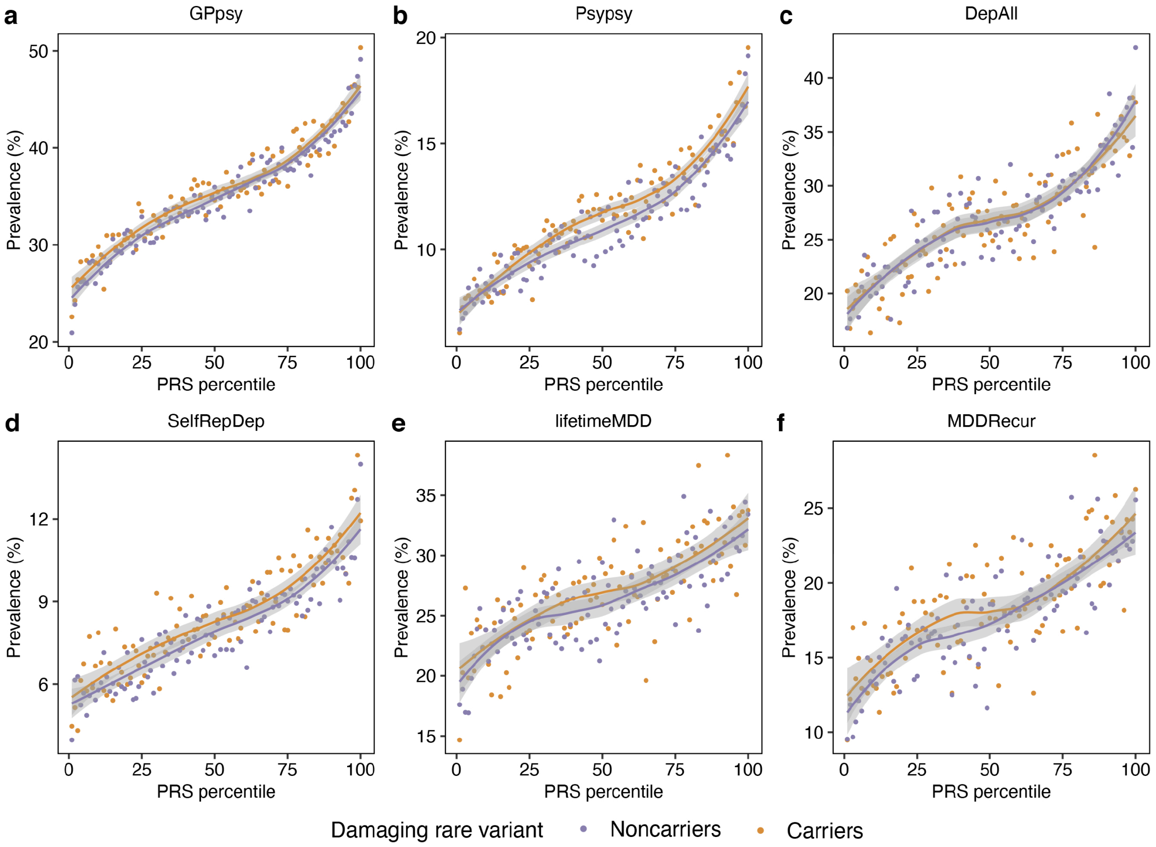


**Figure S9. Prevalence of depression for PRS percentile in PTV or damaging missense variant carriers and noncarriers.** (**a**) GPpsy, (**b**) Psypsy, (**c**) DepAll, (**d**) SelfRepDep, (**e**) lifetimeMDD, (**f**) MDDRecur. Each dot represents a sample. The lines are fitted loess regression for carriers and noncarriers separately and grey shadow corresponds to 95% confidence interval of the fit.

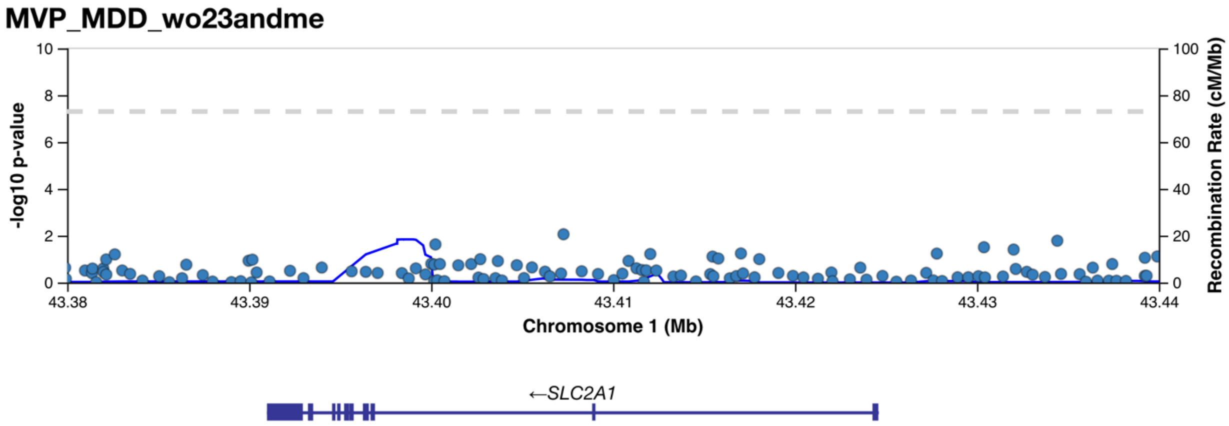


**Figure S10. Regional plot of major depressive disorder (MDD) GWAS[2] *SLC2A1* locus.** The GWAS association p-value for MDD were plotted against genomic position.
